# Risk Factor Profile in Different Subtypes of Acute Ischemic Stroke: A Prospective Observational Study

**DOI:** 10.1101/2023.11.24.23299009

**Authors:** Rajkamal Chaudhari, Amrita Parikh, Yagnya Dalal, Dev Desai, Ami Parikh

## Abstract

**Introduction:** Acute ischemic stroke is a major subtype of acute stroke. More than 65% of stroke-related deaths occur in developing countries. Various modifiable and non-modifiable risk factors are associated with the stroke. TOAST classification categorizes ischemic stroke into five etiologic subgroups.

**Aims:** To identify subtypes of acute ischemic stroke and compare the influence of various risk factors on them.

**Methodology:** A prospective observational study was conducted in a tertiary care teaching hospital. 200 consecutive patients diagnosed as having acute ischemic stroke were randomly enrolled. Patients were categorized into 3 groups-Group A: 18-39 years (17%); Group B: 40-60 years (20%) and Group C: >60 years (63%). Stroke subtypes were ascertained using TOAST classification. Data analysis was done. For categorical variables, data values were represented as numbers and percentages. A chi-square test was applied to find the level of significance. p value<0.05 was considered significant.

**Results:** The study included 136 men and 64 women. Overall male dominance (Male: Female = 2.125:1), while in Group A, strong female dominance (Male: Female = 1:4.67) was observed. The commonest subtype was embolism (29.4%) in Group A, and small vessel disease in Group B, and C ((30% & 46.03% respectively). Hypertension was the commonest risk factor (62%). A higher incidence of hypertension was found in Group A (73.02%, p = 0.002), dyslipidemia in Group B (45%, p = 0.004), valvular heart disease (29.4%, p = 0.00001), and atrial fibrillation (29.4%, p = 0.005) in Group-C. Smoking and diabetes mellitus were strongly related to the male gender (p = 0.000001, p=0.002 respectively) and valvular heart disease to the female gender (p= 0.0002).

**Conclusions:** Awareness of risk factors and lifestyle modifications may have a bearing on stroke prevention. Cardiovascular risk factors in young patients mandate the need for robust prevention and screening.

## INTRODUCTION

Acute ischemic stroke – a major subtype of acute stroke occurs due to loss of blood supply to a part of the brain which initiates an ischemic cascade due to free radical production and damage to the endothelial lining. The global burden of stroke continues to rise mainly in developing countries despite of decrease in the incidence and mortality of stroke in the last 20 years [1].

Stroke is the third main cause of mortality and a significant cause of disability in survivors. More than 65% of stroke-related deaths occur in developing countries [2]. Various modifiable and non-modifiable risk factors are associated with the stroke. Non-modifiable risk factors include age, sex, ethnicity, race, and heredity. Modifiable risk factors include, but are not limited to hypertension, diabetes mellitus, dyslipidemia, atrial fibrillation, smoking, and alcoholism [3].

Treatment options are limited to thrombolysis, but only a few patients receive it due to constraints in its application time and indication[4]. So preventive measures including lifestyle modifications remain the most important strategy to reduce the stroke burden [5]. Ischemic stroke has different subtypes based on etiologies. TOAST classification was introduced in 1993 to improve the sub-classification of ischemic stroke[6]. TOAST classification categorizes patients with ischemic stroke into five subgroups according to the presumed etiological mechanism. They are classified as large artery atherosclerosis (LAA), embolic, small vessel disease (SVD), stroke of undetermined etiologies, and other determined etiologies [7]. There are limited studies on stroke subtypes in India because many health centers don’t have the facilities for magnetic resonance imaging (MRI) and Doppler scans. Often, where the facilities are available, they are unaffordable [8].

## METHODOLOGY

A prospective observational study was conducted in a tertiary care teaching hospital after approval from IRB. 200 consecutive patients diagnosed as having acute ischemic stroke on CT scan or MRI were randomly enrolled in the study after obtaining written consent. Exclusion criteria were age< 18 years, patients with stroke due to intracranial hemorrhage, neoplasms, or CNS infections. Patient’s history, demographic details, clinical findings, and laboratory and radiological reports were entered in the Performa designed to classify the stroke and evaluation of risk factors. As per JNC-8 guidelines, hypertension was classified as stage-1 (systolic BP between 140-159, diastolic BP between 90-99 mm Hg), stage-2 (systolic BP> 160, diastolic BP>=100 mm Hg). Patients who were not previously diagnosed as hypertensive or not on any antihypertensive medication, but had increased blood pressure at the time of presentation due to Cushing’s reflex were not considered hypertensive then. These patients were considered hypertensive if they showed blood pressure findings of hypertension according to JNC-8 guidelines after 7 days of acute ischemic stroke. Patients were considered diabetic when random blood sugar >200 mg/dl glycosylated Hb>6.5 or fasting blood sugar>126 mg/dl and postprandial blood sugar>200 mg/dl. A person who smoked 100 or more bidis or cigarettes during his/her lifetime was considered a smoker. The patient was considered to be dyslipidemic when he/she was already diagnosed as having dyslipidemia, on anti-cholesterol drugs, total cholesterol>200 during fasting, triglycerides>150, LDL>100, HDL<40 in current blood investigation. The patient was said to have cardiovascular disease when he/she had a history and investigatory evidence of coronary artery disease or its complications.

Patients were categorized into 3 groups. They were-

1. 18-39 years (Group-A)
2. 40-60 years (Group-B)
3. >60 years (Group-C)

All patients underwent biochemical investigations like complete blood count, Blood sugar, glycosylated Hb, serum creatinine, blood urea, serum electrolytes, PT with INR, SGPT, fasting lipid profile, and cardiac biomarkers. ANA, APLA, and tests for hypercoagulable states were prescribed when indicated. ECG and echocardiography were done in all patients. All patients underwent a CT scan and MRI of the brain along with MR angiography for extracranial and intracranial vessels.

Subtypes of stroke were ascertained using TOAST classification-

1. Large vessel atherosclerosis
2. Embolism
3. Small vessel disease
4. Stroke of undetermined etiologies
5. Other determined etiologies

### Sample size calculation

The prevalence of Acute ischemic stroke in India is 1.05 – 1.5 per 100 persons[9]

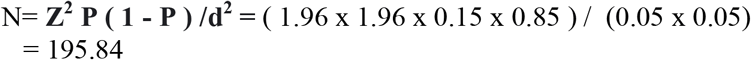

Data values were entered into MS Excel and statistical analysis was done by using IBM SPSS Version 26.0. For categorical variables, data values were represented as numbers and percentages. The chi-square test was applied to find the level of significance and p value<0.05 was considered significant.

## RESULT

Out of a total of 200 patients, 34 belonged to Group A, 40 to Group B, and 126 to Group C. Mean age was 61.08 (SD ±11.63) years. In our study, overall male predominance was noted with 68% (n=136). Female patients were 32% (n=64) in the study. The overall ratio of Males to females was 2.125:1. Female predominance was noted in Group A (n=34) with 82.35% as compared to males (17.65%). In Group B (n=40), males and females were in equal numbers. In Group C (n=126), males outnumbered (87.3%) the female (12.7%) patients. The ratio of Males to females in Group-A was 1:4.6, in Group-B was 1:1, and in Group-C was 6.9:1. (Table 1)

**Table 1:** Age and Gender distribution in different age groups.

| Age (years) | Male | Female | Total |
| --- | --- | --- | --- |
| 18-39 (Group-A) | 06 | 28 | 34 |
| 40-60 (Group-B) | 20 | 20 | 40 |
| >60 (Group-C) | 110 | 16 | 126 |
| Total | 136 | 64 | 200 |

In our study, hypertension emerged as the commonest risk factor in 62% of patients with the maximum number of patients in Group C (73.02%), followed by smoking (47%) with maximum numbers in Group A (52.9%), diabetes mellitus (28%) with maximum numbers of patients in Group-C (33.33%), dyslipidemia (24%) with maximum numbers of patients in Group-B (45%), atrial fibrillation (14%) and valvular heart disease (8%) with a maximum number of patients in Group-A (29.4% each). Hypertension was common in the older age group (p-value 0.018), while atrial fibrillation and valvular heart diseases were common in the younger age group (p-value 0.046 and 0.00002 respectively). (Table 2)

**Table 2:** Age group-wise comparison of risk factors.

| Risk factors | No. of patients | Group-A (18-39 years) n =34 | Group-B (40-60 years) n=40 | Group- C >60 years n=126 | p-value |
| --- | --- | --- | --- | --- | --- |
| Hypertension | 124 | 08(23.5%) | 24 (60%) | 92 (73.02%) | 0.002 |
| Smoking | 94 | 18 (52.9%) | 14 (35%) | 62 (49.2%) | 0.218 |
| Diabetes mellitus | 56 | 06 (17.64%) | 08 (20%) | 42 (33.33%) | 0.202 |
| Dyslipidemia | 48 | 04 (11.76%) | 18 (45%) | 26 (20.63%) | 0.004 |
| Atrial fibrillation | 28 | 10 (29.4%) | 06 (15%) | 12 (9.5%) | 0.005 |
| Valvular heart disease | 16 | 10 (29.4%) | 04 (10%) | 02 (1.6%) | 0.00001 |

Out of 124 hypertensive patients; 48 patients (39%) had hypertension for more than 10 years. Hypertension was detected for the first time in 8 patients (80%) in Group A, 14 patients (58.3%) in Group B and 40 patients in Group C (45.45%). Out of 56 diabetic patients, 36 (64.3%) had diabetes for more than 10 years duration. Out of 48 patients with dyslipidemia; 67% had raised LDL, followed by raised cholesterol in 42%, low HDL in 33%, and raised triglycerides in 21% of patients. The younger age group (Group A) was more affected by atrial fibrillation and valvular heart disease.

Out of 94 smokers; 92 (97.9%) were males and 2 (2.1%) were females. Out of 124 hypertensive patients, 80 were males and 44 were females. No significant gender preponderance was noted in patients with diabetes, dyslipidemia, and atrial fibrillation. Valvular heart disease was more common in females (n=12) as compared to males (n=4). (Table 3)

**Table 3:** Gender-wise comparison of risk factors.

| Risk factors | Male<br>n=136 | Female<br>n=64 | p-value |
| --- | --- | --- | --- |
| Hypertension | 80 | 44 | 0.431 |
| Smoking | 92 | 02 | 0.000001 |
| Diabetes mellitus | 28 | 28 | 0.002 |
| Dyslipidemia | 26 | 22 | 0.0388 |
| Atrial fibrillation | 16 | 12 | 0.237 |
| Valvular heart disease | 04 | 12 | 0.0002 |

The commonest etiological factor for acute ischemic stroke was small vessel disease (39%), followed by LAA stroke (Large artery atherosclerosis in 23.5%), cardio-embolism (21.5%), and undetermined etiology (10.5%). Other determined causes comprised 5.5%. (Table 4)

**Table 4:** Etiology-wise distribution of patients.

| Etiology | Number of patients | % of patients |
| --- | --- | --- |
| Small vessel disease | 78 | 39% |
| LAA (Large artery atherosclerosis) | 47 | 23.5% |
| Embolism | 43 | 21.5% |
| Undetermined etiology | 21 | 10.5% |
| Other determined etiology | 11 | 5.5% |
| Total | 200 | 100% |

Correlation was done between risk factors and subtypes of acute ischemic stroke. It was found in the study that hypertension was the most common risk factor across all subtypes of ischemic stroke. A strong correlation was noted between the embolic subtype of ischemic stroke and atrial fibrillation as well as valvular heart disease as risk factors. (Table 5)

**Table 5:**
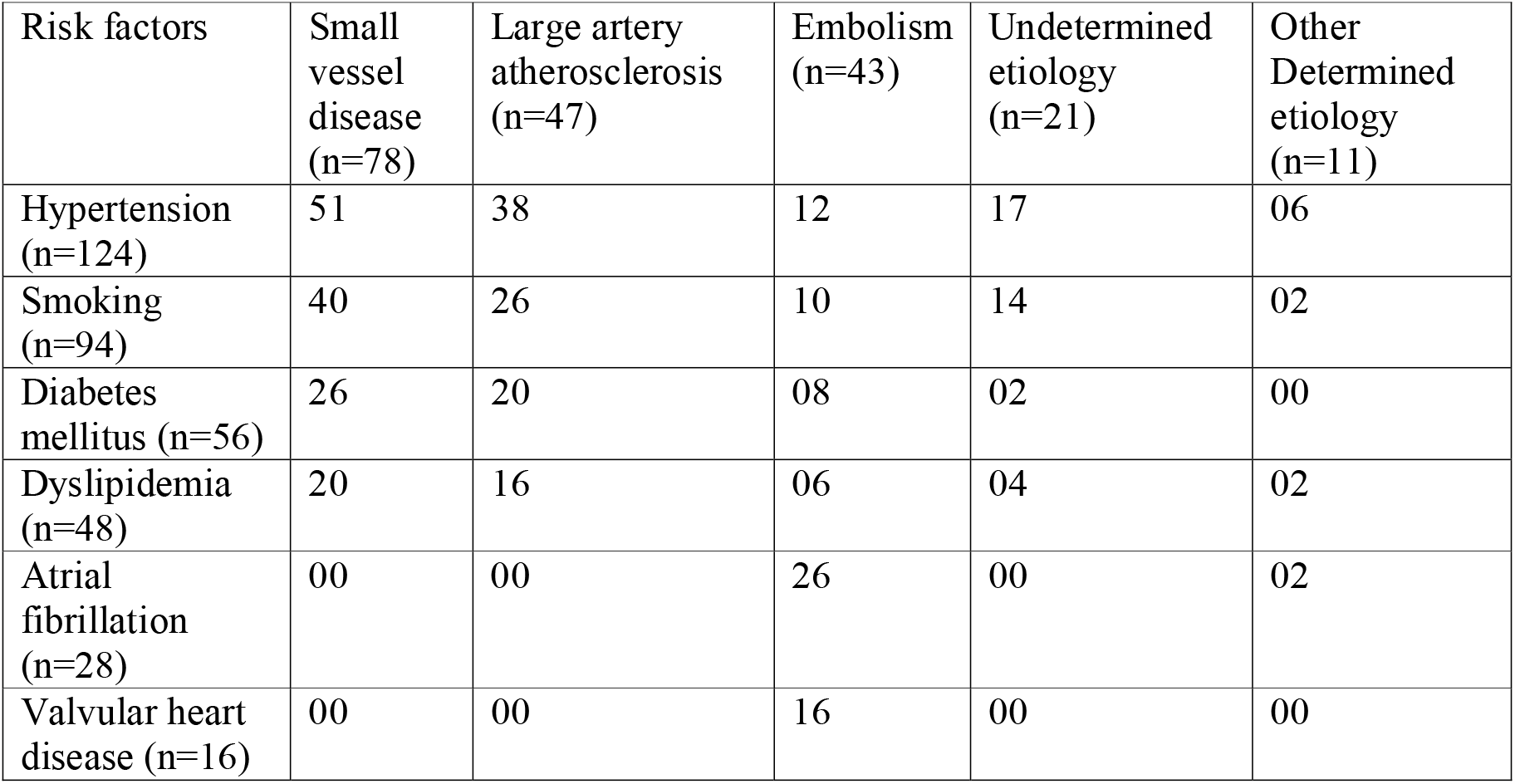
Correlation of Subtypes of acute ischemic stroke to risk factors.

Age groups of the patients were correlated with subtypes of acute ischemic stroke. Embolism was found in 29.4% of patients in Group A, followed by small vessel disease (23.5%), stroke due to undetermined etiologies and stroke due to other determined causes (17.64% each), and large artery atherosclerosis (11.76%). In Group B, small vessel disease and large artery atherosclerosis were of almost equal distribution (30% and 27.5% respectively), followed by cardio-embolism (17.5%), stroke due to undetermined etiology, and other determined etiology contributed 12.5% each. Small vessel disease was the most common subtype in group C (46.03%), followed by large artery atherosclerosis (25.4%), embolism (20.63%), and undetermined etiology (7.9%). (Table 6)

**Table 6:** Age-wise distribution of subtypes of ischemic stroke.

| Age group | Group-A (18-39 years)n=34 | Group-B (40-60 years)n=40 | Group-C (>60 years) n=126 | p-value |
| --- | --- | --- | --- | --- |
| Small vessel disease (n=78) | 08 (23.5%) | 12 (30%) | 58 (46.03%) | 0.024 |
| Large artery atherosclerosis (n=47) | 04 (11.76%) | 11 (27.5%) | 32 (25.4%) | 0.200 |
| Embolism (n=43) | 10 (29.4%) | 07 (17.5%) | 26 (20.63%) | 0.428 |
| Undetermined etiology (n=21) | 06 (17.64%) | 05 (12.5%) | 10 (7.9%) | 0.234 |
| Other determined etiology (n=11) | 06 (17.64%) | 05 (12.5%) | 00 | - |

## DISCUSSION

Stroke is a major public health burden with significant morbidities and mortalities. Stroke predominantly occurs in males in the late years of life. There are modifiable and non-modifiable risk factors for stroke. The prevalence of risk factors varies in different populations and different age groups. The youngest patient was 18 years and the oldest was 76 years in the present study. The stroke prevalence is high in the age group of >60 years (n=126). The mean age of our study was 61.08 years. Our study correlates with the studies done by Renjen PN et al[10]and Vaidya CV et al[11] with mean ages of 57.1 years and 60.2 years respectively.

In our study, overall male preponderance was found (68%) which correlates with studies done by Manorenj S et al[12]and Renjen PN et al[10] with a male preponderance of 65.4% and 67.6% respectively. Female predominance was found in the present study in the younger age group (Group-A: <40 years) with 82.35% of patients being female. This correlates with a study conducted by Ekker MS et al[13].

In our study; the frequency of small vessel disease was highest (39%), followed by large-artery atherosclerosis (LAA) (23.5%), embolism (21.5%), stroke of undetermined etiologies (10.5%) and other determined causes (5.5%). Other determined causes were Takayasu’s arteritis, tubercular arteritis, Moyamoya disease, fibromuscular dysplasia, and APLA. Our study showed that small vessel disease was most common in older age (>60 years) (46.03%). Stroke due to large artery atherosclerosis was more common in the age group of 40-60 years (27.5%). Embolic stroke, stroke due to undetermined etiology, and stroke due to other determined etiology were more common in young patients (<40 years) (29.4%, 17.64%, 17.64% respectively). It was found in the study that hypertension was the most common risk factor across all subtypes of ischemic stroke. A strong correlation was noted between the embolic subtype of ischemic stroke and atrial fibrillation as well as valvular heart disease as risk factors.

In the present study, hypertension was found to be the most prevalent vascular risk factor (62%) across all subtypes of stroke, followed by smoking (47%), diabetes mellitus (28%), dyslipidemia (24%), atrial fibrillation (14%) and valvular heart disease (8%). In a study conducted by Zafar A et al, systemic hypertension emerged as a single risk factor associated with stroke[7]. Hypertension was the most frequently identified risk factor in all subtypes of ischemic stroke except for stroke of other determined etiology, which is consistent with the published literature[14]. Hypertension causes small vessel disease (SVD) by changing brain vasculature through fibrinoid deposition in the small vessel and hypertrophy of smooth muscle, thus leading to brain ischemia in affected areas [15]. Another condition thought to be associated with SVD and undetermined etiology subtype is nocturnal hypertension which is affected by circadian rhythm. Nocturnal blood pressure variability may be associated with wake-up stroke incidence which occurs in 20% to 30% of ischemic stroke[16]. Hypertension was detected for the first time in 42(33.8%) patients (n=124). Detection of hypertension at the time of stroke for the first time in all age groups mandates early screening programs in the general population.

Diabetes mellitus is a well-known risk factor that causes both microangiopathy and macroangiopathy. The pathophysiological changes of diabetic cerebral vessels differ in comparison with non-diabetic patients. Diabetes mellitus predisposes to small vessel lacunar infarcts [17]. The present study reveals that 28% of patients had diabetes. 36 (64.3%) patients had diabetes for more than 10 years duration (n=56). Diabetes was more common in male patients (p=0.002) in our study. Smoking appears as an important risk factor for acute ischemic stroke in our study. 47% were smokers among the patients studied. This correlates well with Renjen PN et al[10](39% smokers). Smoking was a major risk factor in small vessel disease and large artery atherosclerosis as well as other subtypes of ischemic shock in our study. Smoking was strongly related to male patients (p-value 0.000001) in our study. In the present study, dyslipidemia was noted in 24% of patients, which correlates with a prior study done by Zafar A et al[7]. Dyslipidemia was a major risk factor in small vessel disease, large artery atherosclerosis, and embolism. Dyslipidemia in the form of high total and LDL cholesterol is an important risk factor in the development of ischemic stroke [18]. Atrial fibrillation and valvular heart disease were less common risk factors found in our study which correlated with a study conducted by Aquil et al[19]. Atrial fibrillation (AF) is a disease of the elderly. However, in the Indian subcontinent, valvular heart disease contributes significantly to the development of valvular AF in the younger age group (p=0.005). All 10 patients with valvular heart disease in Group A developed atrial fibrillation in our study. Atrial fibrillation and valvular heart disease were present in patients with embolic subtypes of stroke in our study. In our study, valvular heart disease was more common in female patients (p=0.0002). The presence of valvular heart disease and atrial fibrillation in the age group<60 years indicates their burden as risk factors in acute ischemic stroke in the younger population.

Hypertension was the commonest risk factor in Group C (p=0.002). Though diabetes was commoner in Group C (33.33%), the younger age groups were also affected. Smoking was common across all age groups. 45% of Group-B patients had dyslipidemia. The commonest subtype of stroke in Group A was embolism, and in Group B were SVD and LAA. While the commonest subtype in Group C was SVD.

## CONCLUSION

The influence of risk factors is different at different ages. Awareness of these risk factors before the occurrence of stroke is crucial to primary prevention. Identification of one or more risk factors in patients with acute ischemic stroke is also the cornerstone of secondary prevention. Cardiovascular and valvular risk factors in our study group of young patients mandate the need for robust prevention as well as screening strategies.

## Data Availability

All data produced in the present study are available upon reasonable request to the authors

